# RANDOMIZED CONTROL STUDY TO ANALYSE THE EFFICACY BUDESONIDE NASAL DOUCHING ON ALLERGIC FUNGAL RHINOSINUSITIS

**DOI:** 10.1101/2024.05.02.24306490

**Authors:** Sowparnika TR, Ramaneeshwaran, Ashu Seith Bhalla, Karan Madan, Yashdeep Gupta, Rupesh K Srivastava, Ganesh Kumar Viswanathan, Gagandeep Singh, Alok Thakar, Rakesh Kumar, Kapil Sikka, Shuchita Singh Pachaury, Hitesh Verma

**Affiliations:** Department of Otorhinolaryngology and Head and Neck Surgery, AIIMS New Delhi; Department of Radiology, AIIMS New Delhi; Department of Pulmonology and Sleep Medicine, AIIMS New Delhi; Department of Endocrinology and Metabolism, AIIMS New Delhi; Department of Biotechnology, AIIMS New Delhi; Department of Hematology, AIIMS New Delhi; Department of Microbiology, AIIMS New Delhi; Department of Otorhinolaryngology and Head and Neck Surgery

## Abstract

**INTRODUCTION:** Allergic Fungal Rhinosinusitis is a hypersensitivity against fungal antigens. Primary surgical management is complimented with medical management in preoperative and postoperative phases for better intraoperative field & to limit the recurrence. The effect of systemic steroids & itraconazole showed promising outcomes as a solo agent or in combination under preoperative status. Recent studies showed auspicious conclusions with steroid-based nasal douches. The current study aims to assess the effectiveness of medical management in combination with steroid-based nasal douches.

**METHODOLOGY:** This prospective study will recruit 40 clinico-radiological proven cases of AFRS after ethical approval. The study population will be randomized into two groups based on a computer program. Group A will receive systemic itraconazole and steroids & Group B will receive additional budesonide nasal douching. The outcomes will be studied clinically by SNOT-22 scoring system & Meltzer nasal polyp scoring system, biochemically with absolute eosinophil counts, aspergillus specific IgE and total IgE, radiologically with Lund-Mackay system and 20-point CT scoring system after 6 weeks of initiation of the treatment.

**ETHICS AND DISSEMINATION:** We received an AIIMS ethical board. Ref no-AIIMSA00641 dated 21.03.24 & CTRI (Clinical Trial Registry India) acknowledgement number - REF/2024/03/081566 dated 29. 03.24. We will follow the ethical committee protocol and instructions received from reviewers.

**STRENGTHS:**

1. AFRS is a bothersome illness both for treating doctors and patients. This study is a pilot study on a combination of past and recently proven medical therapies for AFRS.
2. The study will generate data on new or complete medical management for AFRS.
3. The duration of planned treatment is 6 weeks. The study will generate data for future studies where prolonged medical treatment can be planned.

**LIMITATIONS:**

1. This will be a single-centred study. The validation for the expected outcome will require a multicentric study.
2. The study topic is planned for an MS student thesis so the follow-up period will be 3-6 months.

## INTRODUCTION

### Description of condition

Allergic fungal rhinosinusitis (AFRS) is a hypersensitivity against fungal antigens. The diagnosis is established with characteristic clinical, radiographic, and histopathologic findings. Till now, the disease treatment methods are understood incompletely as they require both surgical & medical management. The couple of medical management in isolation & combination showed enlightening outcomes, but none of the studies showed complete effectiveness.

### Description of intervention

The most followed treatment for AFRS is surgery, which is predominantly endoscopic clearance of fungal muck, polyps & allergic mucin. Given persistent causes, recurrences are not infrequent and require adjunct medical therapies to minimize recurrences, and disease load & to improve quality of life. Studies have shown enlightening results of systemic steroid, systemic antifungal {1} and steroid nasal douching in pre-surgical settings as solo agents & in combination preoperatively. This work aims to improve medical therapy by combining older with recent treatment methods by assessing the outcome of a combination of systemic therapies with topical steroids. Treatment response will be assessed with clinical-radiological and biochemical parameters as per Bent & Kuhn criteria{2} This will be the first study which evaluates the efficacy of pre-surgical steroid rinses with systemic therapies.

### How intervention might work

Fungal hypersensitivity is managed by both steroid & antifungal therapy. Both drugs reduce inflammation, fungal load and hyper-sensitivity which reduces the disease burden{3}. The combination of systemic therapies with topical irrigation will work more efficiently in comparison to solo agents.

### Existing knowledge

Allergic fungal rhinosinusitis (AFRS) indeed stands out as a distinct entity among sinus diseases, characterized by its non-invasive nature and association with fungal antigens hypersensitivity{4,5}. Its unique clinical presentation, histopathological features, and radiological findings set it apart from other forms of sinusitis. Typically affecting younger individuals, with a mean age range of 21–33 years, AFRS requires careful management to alleviate symptoms and prevent recurrence. The cornerstone of AFRS management lies in surgical intervention aimed at removing polyps, fungal debris, and allergic mucin from the sinuses{6}. Preoperative medical therapy plays a crucial role in preparing the patient for surgery by reducing the extent of disease, enhancing the surgical field, and minimizing potential complications. Postoperative medical therapy is equally important in preventing recurrence{7}. Both therapy’s primary objective is to maintain sinus health and suppress inflammatory responses that could lead to the re-emergence of symptoms. In the preoperative management of allergic fungal rhinosinusitis (AFRS), systemic steroids and systemic antifungals {8–11} are commonly employed as medical therapies, with or without topical steroids.

Topical steroids have not shown a significant impact on disease load when used in isolation, they reduce the level of local inflammation which indeed improves symptoms, potentially optimizing the surgical field and enhancing the effectiveness of surgical intervention. The combination of topical steroid spray with systemic steroids does indeed appear to be a commonly utilized approach in managing allergic fungal rhinosinusitis (AFRS) as topical therapy can be given for a prolonged period. Systemic steroid therapy has certainly demonstrated a significant impact on reducing disease burden in AFRS. Using oral steroids in a tapering dose regimen has been particularly effective in producing both subjective and objective improvements in AFRS symptoms{12}. However, the optimal duration and dosing of systemic steroids remain variable across studies, with no consensus established due to inconsistent outcomes among different regimens{13}. One major challenge in determining the most effective systemic steroid regimen for AFRS is the lack of standardized dosing protocols and treatment duration. Studies have shown varying results, and as a result, there’s a lack of clear guidance on the ideal dosage and duration of systemic steroid therapy for AFRS. Furthermore, the long list of potential side effects associated with systemic steroid use is a significant limitation. These side effects can range from mild to severe and may include weight gain, mood changes, osteoporosis, diabetes, and increased susceptibility to infections. Because of these risks, systemic steroids are typically used for short periods, with careful consideration given to balancing their potential benefits with the potential for adverse effects.

Combining topical steroids nasal douches with oral steroids has shown promising results in recent studies (14,15). Research indicates that this combination therapy can significantly reduce recurrence rates compared to placebo. The study conducted by Osama et al. in 2014 {14} sheds light on the efficacy of budesonide nasal douching in the management of allergic fungal rhinosinusitis (AFRS). Their findings suggest that budesonide nasal douching holds promise as a valuable tool in the treatment of AFRS. Nasal douching with budesonide involves using a saline solution containing budesonide, a corticosteroid medication, to irrigate the nasal passages. This approach allows for direct delivery of the steroid to the affected sinus tissues, where it can help reduce inflammation and alleviate symptoms associated with AFRS. The study demonstrated improvements in symptom severity, reduction in polyp size, and overall enhancement in sinus health among AFRS patients who underwent budesonide nasal douching.

In the study by Barsha et al., conducted during the COVID-19 period, the focus on preoperative administration of systemic itraconazole as a solo agent sheds light on the potential efficacy of systemic antifungal therapy in reducing the burden of AFRS. Systemic itraconazole may have a significant impact on various dimensions of the disease, potentially improving symptoms and preparing patients for surgery, even amidst challenging circumstances{16–18}. Moreover, diving deeper into the review of medical management, the study by Verma et al{19}, which investigated the combination of systemic steroids with itraconazole in a randomized controlled trial, further bolsters the case for combination therapy. The significant impact observed with both solo and combination therapy on disease dimensions underscores the potential benefits of these treatment approaches{19}.

### Need for this trial

Till now, no consensus & effective medical management is available for primary & recurrent AFRS. This study will generate data on effective medical management which allows an effective reduction of disease load & it will also create data on effective pre-surgical management.

### Dose selection

Systemic medical therapy will consist of oral prednisolone 1 mg/kg/day in tapering doses (1 mg/kg for the first 7 days, 0.8 mg/kg from 8^th^ to 14^th^ day, 0.6 mg/kg from 15-22^nd^ day, 0.4 mg/kg from 23-28^th^ day & 0.2 mg/kg from 29^th^ -42^nd^ day of treatment {8}. Tablet Itraconazole will be prescribed 400mg PO BD on the first day & it will be followed by 200 mg/day till 6 weeks {16}. The topical nasal douches will be based on budesonide. 1mg of budesonide suspension will be mixed in 240 ml of normal saline & the subjects will administer 60 ml of prepared solution in each nostril twice daily for 6 weeks {13}.

The subjects will be followed 2 weekly for the effectiveness of therapy & to document induced side effects. The patients will be assessed clinically for side effects. The cortisol level will be assessed on day 7 and day 21. The liver & kidney functions will be assessed every two-week intervals.

At the end of the proposed study period, the subjects will be re-evaluated in a similar way at the time of recruitment. The subjects with persistent disease will be given the choice to continue medical management further or to go for surgery.

## STUDY OBJECTIVES

1. PRIMARY OBJECTIVES: The primary evaluation point will be the change in clinical & radiological scoring.
2. SECONDARY OBJECTIVES: The secondary outcome will be the change in systemic parameters (absolute eosinophil counts, IgE levels, specific IgE levels and interleukin-5 levels).

## TRIAL DESIGN

Trial registered with Institutional Ethics Committee (AIIMS, New Delhi) & CTRI (Clinical Trial Registry India).

1. AIIMS ethical board - Ref no-AIIMSA00641/21.03.24
2. CTRI trial acknowledgement number - REF/2024/03/081566

### Funding

It is a non-funded study. The study will use the existing setup in AIIMS New Delhi.

This study is designed as a randomized controlled, labelled single-centred trial with two parallel groups and a primary endpoint at 6 weeks after starting treatment. Randomization will be performed as computer-based randomization with a 1:1 allocation.

## METHODS

### STUDY SETTING

The current study is planned under the Department of Otorhinolaryngology, Head and Neck Surgery, AIIMS, New Delhi in collaboration with the Department of Radiology, Pulmonology, Endocrinology, Microbiology, Hematology, and the Department of Biotechnology.

The study period will be from January 2024 to December 2025. The ethical & clinical trial approvals are obtained from the AIIMS Ethics Board & CTRI. The diagnosis of AFRS will be based on Bent and Kuhn’s criteria. The study will enroll 40 subjects. The sample size is calculated by observing the existing workload & past studies done in the department.

The subjects will be randomized into two groups by computer-based randomization method. Group A will receive systemic therapies & group B will receive systemic therapies with steroid nasal douches. A baseline blood parameters, nasal endoscopy & radiology will be performed in both groups. The blood investigations will comprise absolute eosinophil count (AEC), IgE levels, specific IgE levels and serum Interleukin-5 levels will be performed. The nasal irrigation sample will be collected for fungal smear & culture. NCCT nose PNS and orbit will be performed to know the sinus involvement. The parameters will be evaluated in the expert departments.

### ELIGIBILITY CRITERIA

A. INCLUSION CRITERIA: We will enrol clinical-radiologically proven AFRS who will be willing to participate in the study. The subjects should be above 14 years of age.
B. EXCLUSION CRITERIA: The exclusion criteria include intolerant cases to medical management, complicated AFRS & subjects with uncontrolled chronic systemic illness (anticipating toxicity of steroid and itraconazole) where medical management will not be allowed.

### INTERVENTIONS

Group A will receive a combination of systemic therapies. The systemic medical therapy will consist of oral prednisolone 1 mg/kg/day in tapering doses (1 mg/kg for the first 7 days, 0.8 mg/kg from 8^th^ to 14^th^ day, 0.6 mg/kg from 15-22^nd^ day, 0.4 mg/kg from 23-28^th^ day & 0.2 mg/kg from 29^th^ -42^nd^ day of treatment {4}. Tablet Itraconazole will be prescribed 400mg PO BD on the first day & it will be followed by 200 mg/day till 6 weeks {5}. Group B will receive medications the same as Group A with additional budesonide nasal douches. The 1mg of budesonide suspension will be mixed in 240 ml of normal saline & the subjects will administer 60 ml of prepared solution in each nostril twice daily for 6 weeks {14}.

The subjects will be followed 2 weekly for the effectiveness of therapy & to document induced side effects. The patients will be assessed clinically for side effects. The cortisol level will be assessed on day 7 and day 21. The liver & kidney functions will be assessed every two-week intervals. In case of any side effects of steroid or antifungal / noncompliance to the treatment medications will be stopped and the patient will be kept under follow-up. Once the parameters reach the baseline treatment can be re-started.

#### PATIENT TIME LINE

Process of recruitment, randomization and work process has been explained as per Annexure 1.

### DATA COLLECTION

The demographic details and symptomatology by SNOT 22 scoring system {20} will be collected and entered in proforma. The clinical assessment will be done endoscopically {21} & radiological assessment will be done with low-dose NCCT {22, 23}. The laboratory investigations like Absolute eosinophil count, serum total IgE, serum specific IgE and serum Interleukin-5 levels will be sent at the time of recruitment. The assessment tools will be reapplied after 6 weeks of therapy. Patients who will not complete the protocol and will not be able to tolerate medical therapy will be excluded from the study. The loss of follow-up rate is expected to be less than 5%.

### ASSIGNMENT OF INTERVENTION

#### ALLOCATION AND SEQUENCE GENERATION

Participants will be randomly assigned to either a control or experiment group with a 1:1 allocation as per a computer-generated randomization schedule. The allocation sequence is concealed. Randomization is implemented by our team with randomization code. It’s an open-label study without blinding.

#### STATISTICAL METHODS

Qualitative data will be reported as number and percentage whereas quantitative data as mean +/- standard deviation. Intergroup comparison will be done with a chi-square test for qualitative variables and a two-sample T-test for quantitative variables. Statistical calculations will be performed using SPSS software ( version 29). The significant threshold is set at p<0.05.

### DATA MONITORING

#### INTERIM ANALYSIS

The study is reviewed every 6 months as per department protocol for suggestions and modifications to the trial. Data monitoring Committee is not required since it is a short-term study and carries only minimal risks with intervention. An interim analysis will be performed once 50% of the sample size is obtained.

#### HARMS

Any adverse events to the treatment will be monitored throughout the study period. Any events occurring after starting the therapy will be considered as adverse events related to the intervention. Expected adverse events could be the progression of disease and altered systemic parameters.

#### ETHICS AND DISSEMINATION

The study got ethical approval. This is an academic study and it is associated with no to minimal risk to the subjects. The patient will enrolled after written informed consent & they will be followed up according to ethical concerns. The therapy will be withheld in patients with side effects & will be followed according to the pre-existing practice. We will only collect data relevant to the study and approved by the Ethics Committee

#### PROTOCOL AMENDMENTS

Any modifications to the protocol which can bring potential benefit and major impact to the study including changing objectives, study design, and sample size will require approval from the Institutional Ethics Committee.

#### CONSENT

The details of the study will be explained using images and videos by the treating team to the patients willing to undergo the study. Patient Information details and Consent forms will be provided in both English and Hindi language. The model consent form and Patient Information Sheet have been attached in the annexure.

#### CONFIDENTIALITY

All the details of the patients are stored in the secured chamber with limited accessibility. Patient information will not be disclosed without permission from the patient. Individual patient information will not be disclosed at the time of publication.

## CONTRIBUTIONS

Dr Hitesh Verma^13^ conceived the study & edited the manuscript. Dr Sowparnika TR ^1^, and Dr Ramaneeshwaran^2^ initiated the study design and prepared the manuscript. Dr Alok Thakar^9^, Dr Rakesh Kumar^10^, Dr Kapil Sikka^11^, and Dr Shuchita Singh Pachaury^1^ ^were^ involved in the implementation & design of the study. Dr Ashu Seith Bhalla^3^ radiological expertise in clinical trial design. Dr Karan Madan^4^, Dr Yashdeep Gupta^5^, Dr Rupesh K Srivastava^6^, Dr Ganesh Kumar Viswanathan^7^, and Dr Gagandeep Singh^8^ are conducting the laboratory analysis of data. All authors contributed to the refinement of the study protocol and approved the final manuscript.

## DECLARATION OF INTERESTS

No conflict of interest is present. ACCESS TO DATA:

All principal investigators are given access to the data during the study.

## ANCILLARY AND POST TRIAL CARE

The patients will be followed, and surgical & medical interventions will be given whenever necessary.

## DISSEMINATION POLICY-TRIAL RESULTS

The study will be published with results in critically acclaimed journals.

## Data Availability

All data produced in the present study are available upon reasonable request to the authors

## >ANNEXURE 1

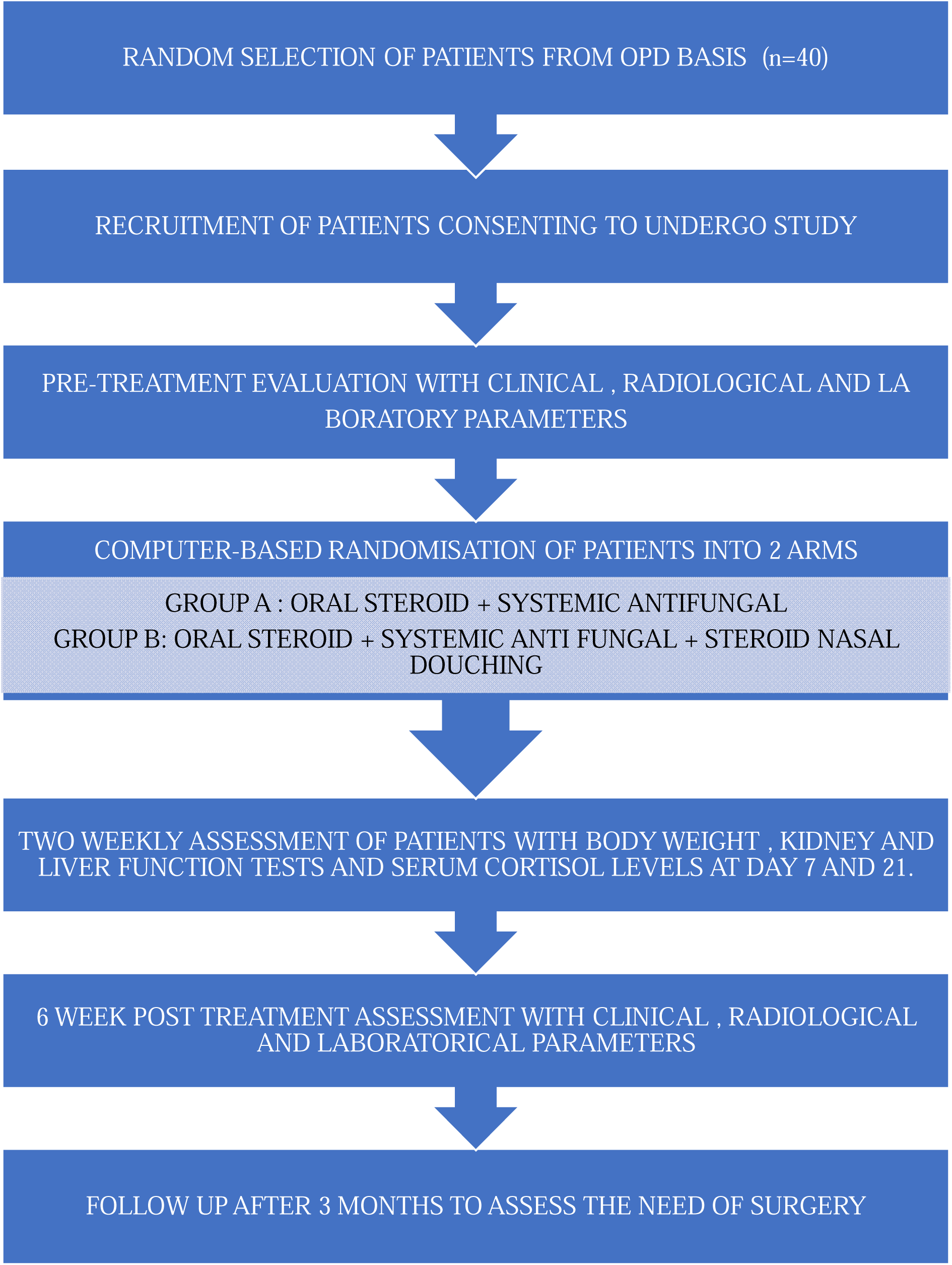

## ANNEXURE 2: MODEL PROFORMA SHEET

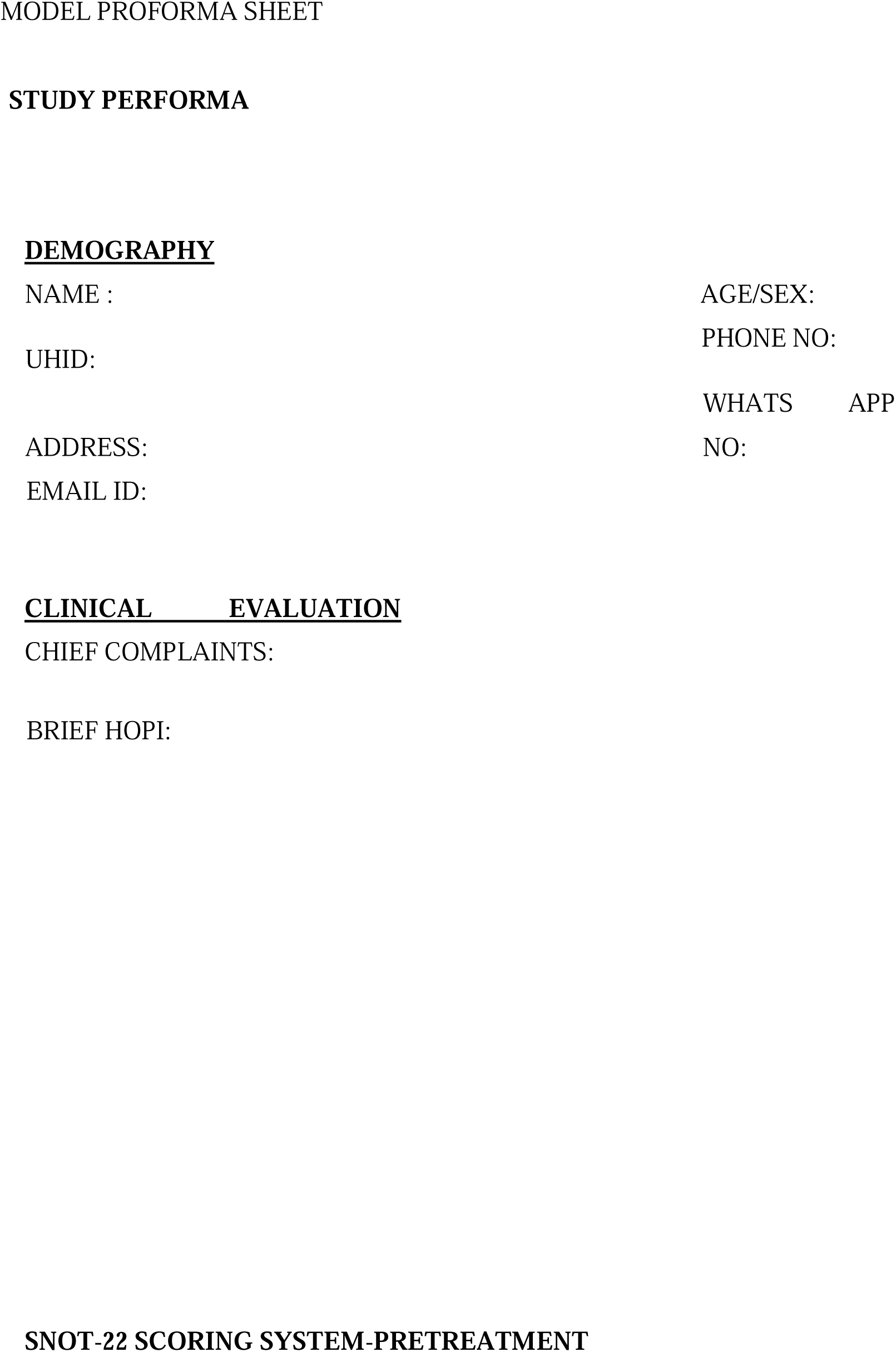
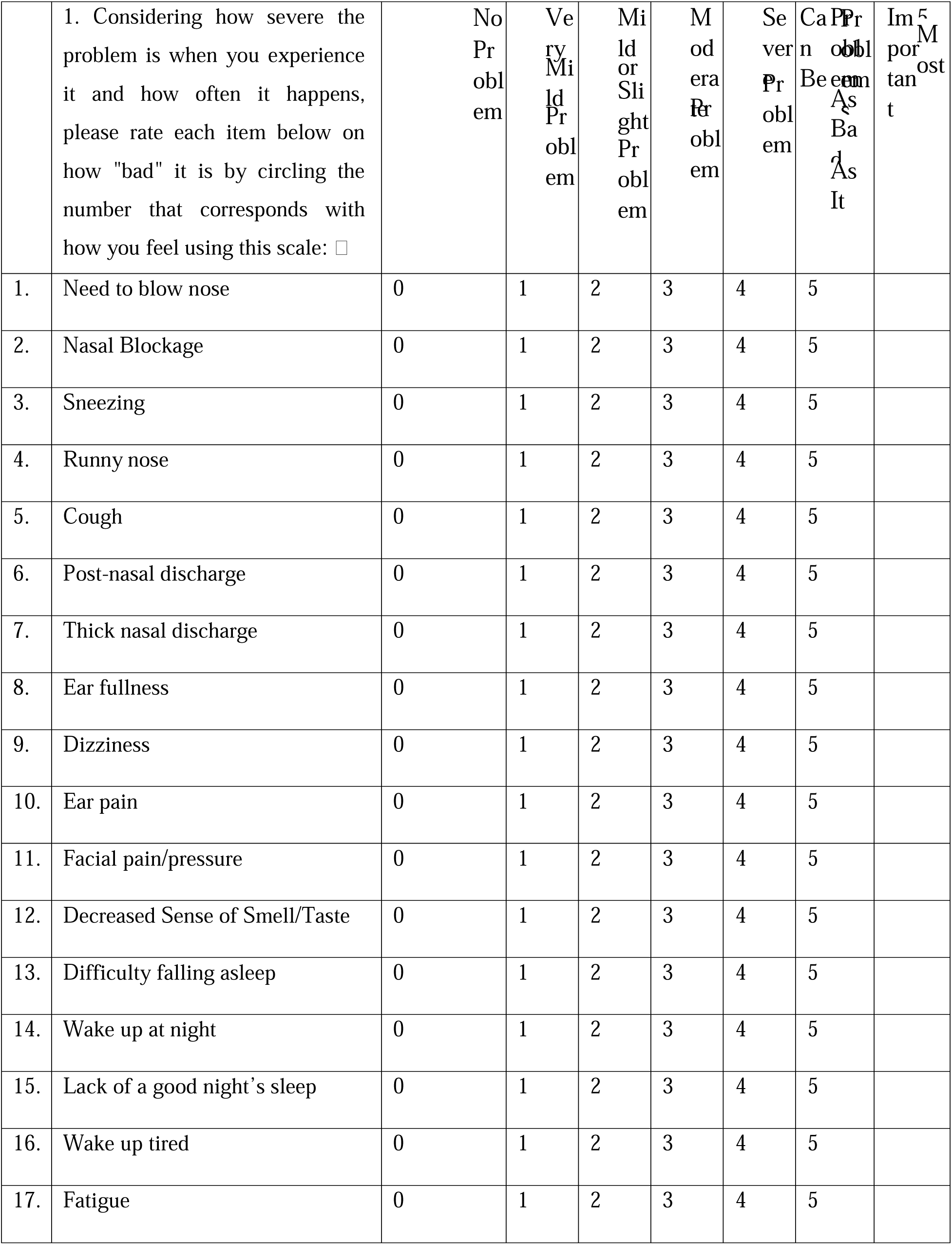
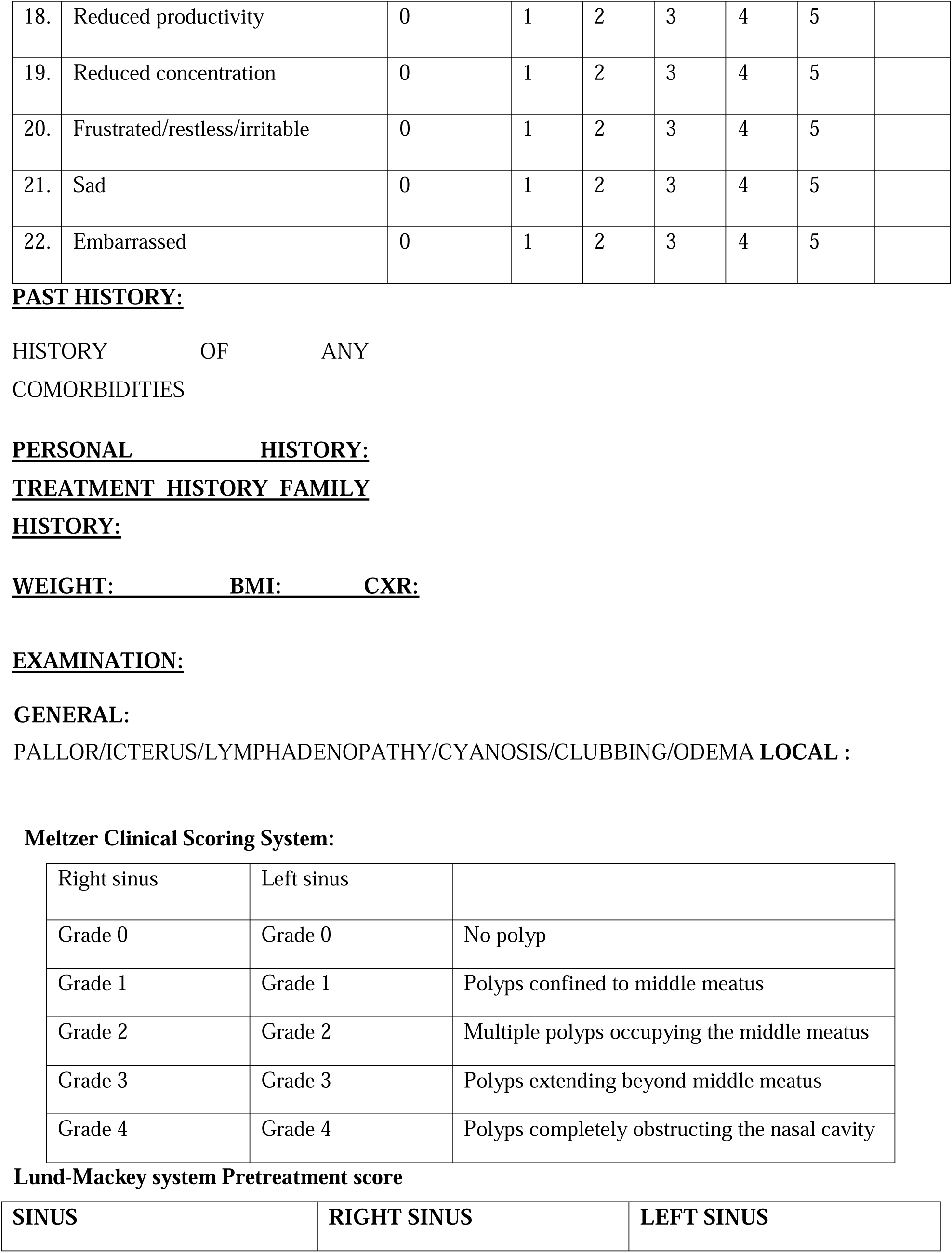
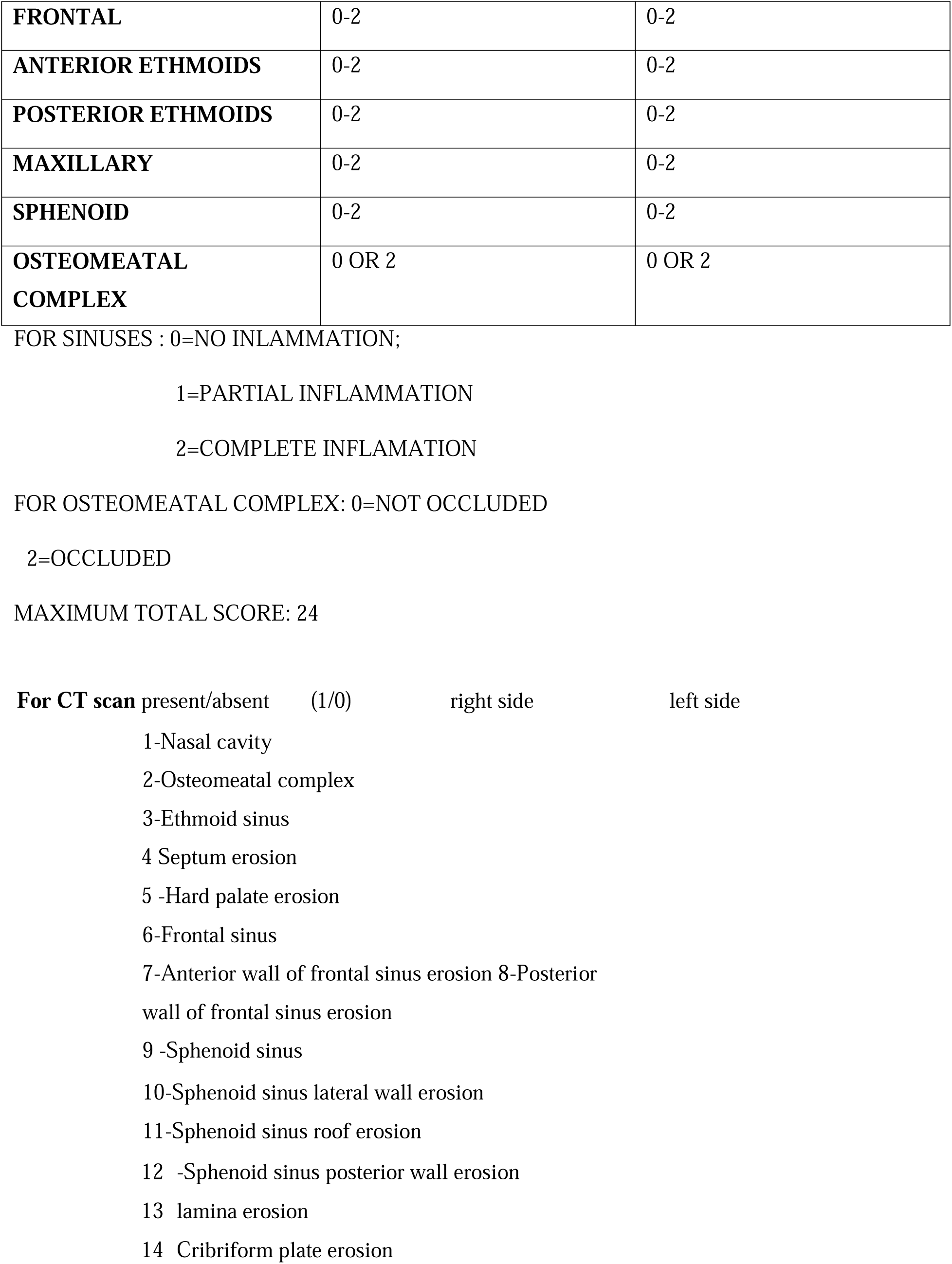
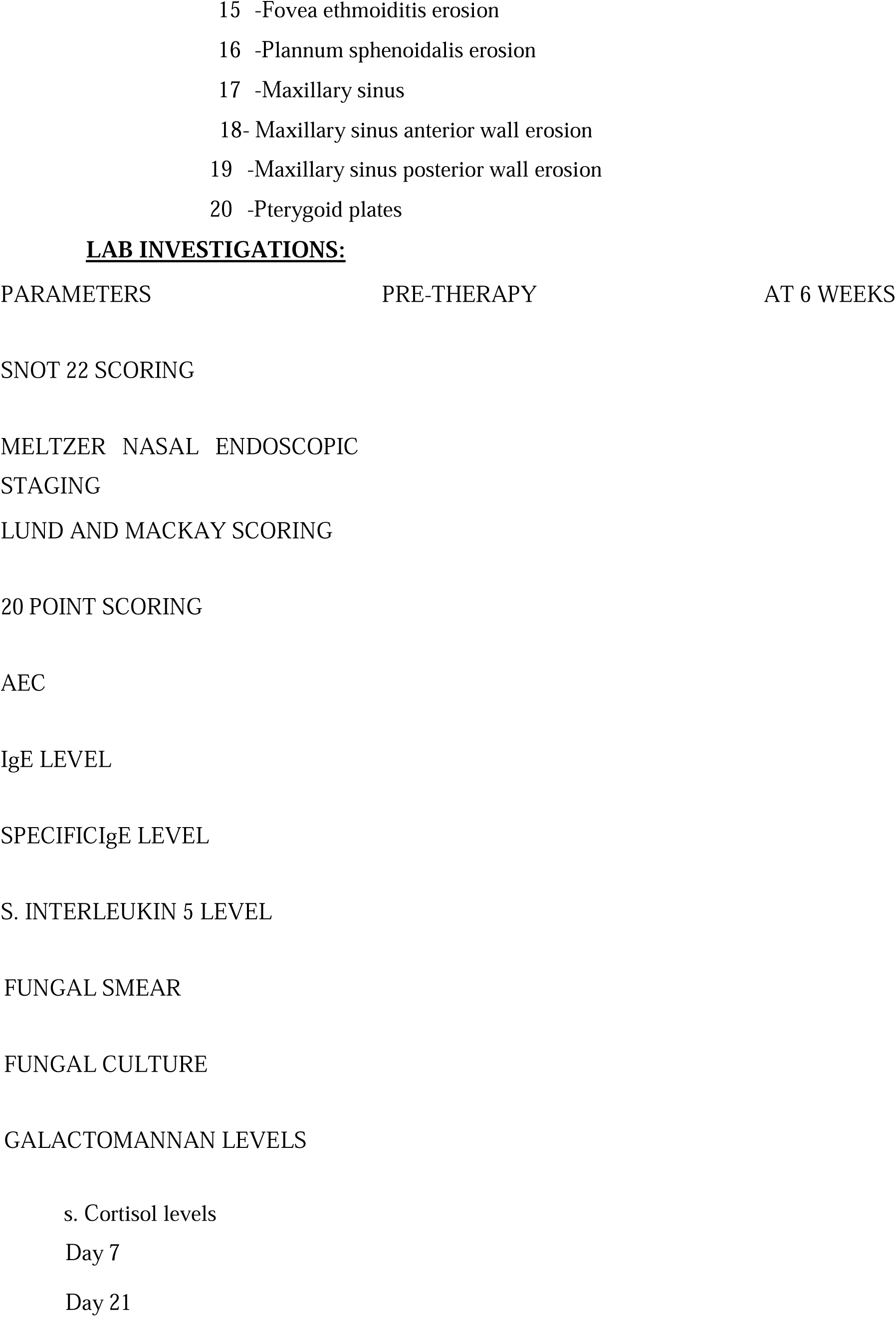
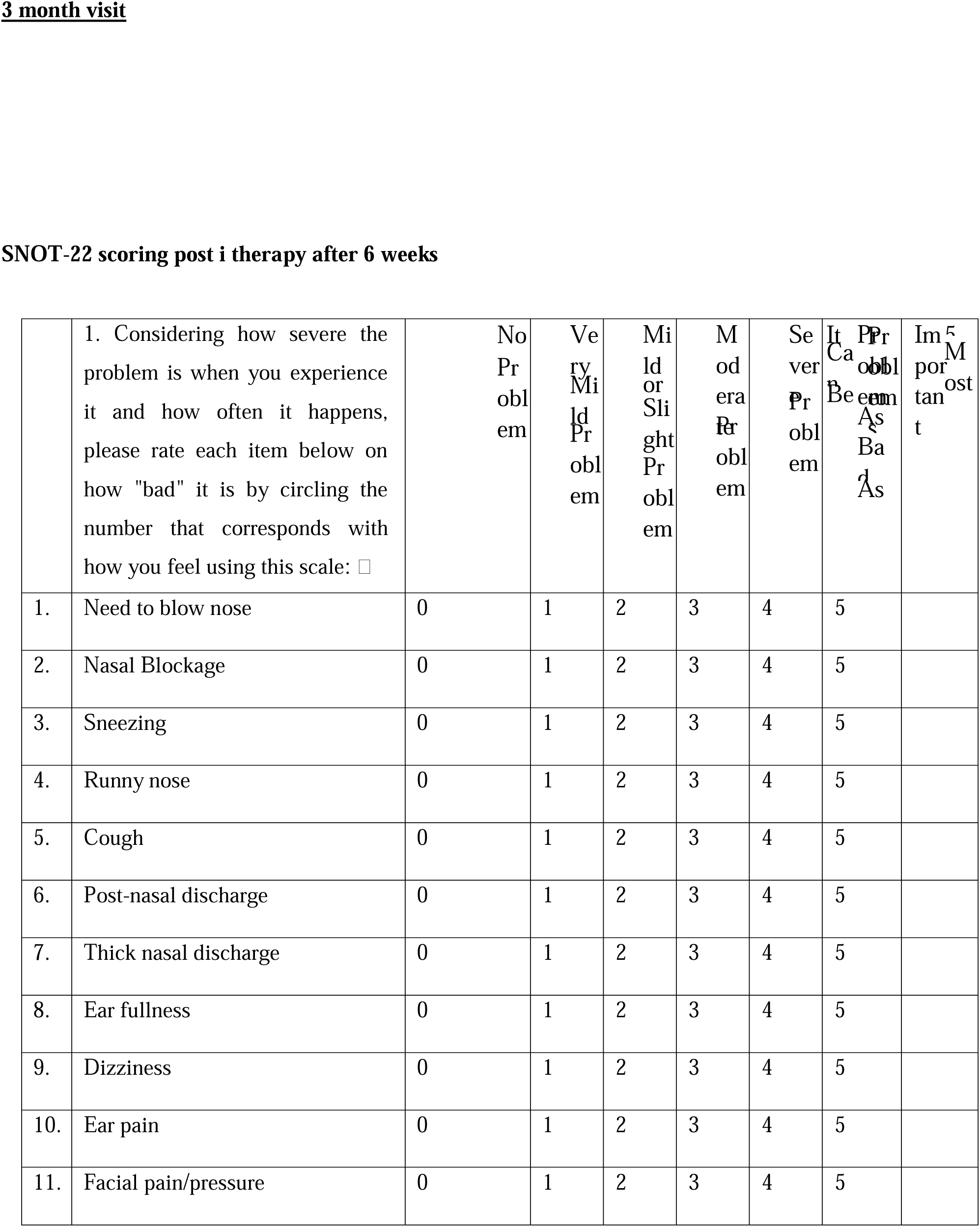
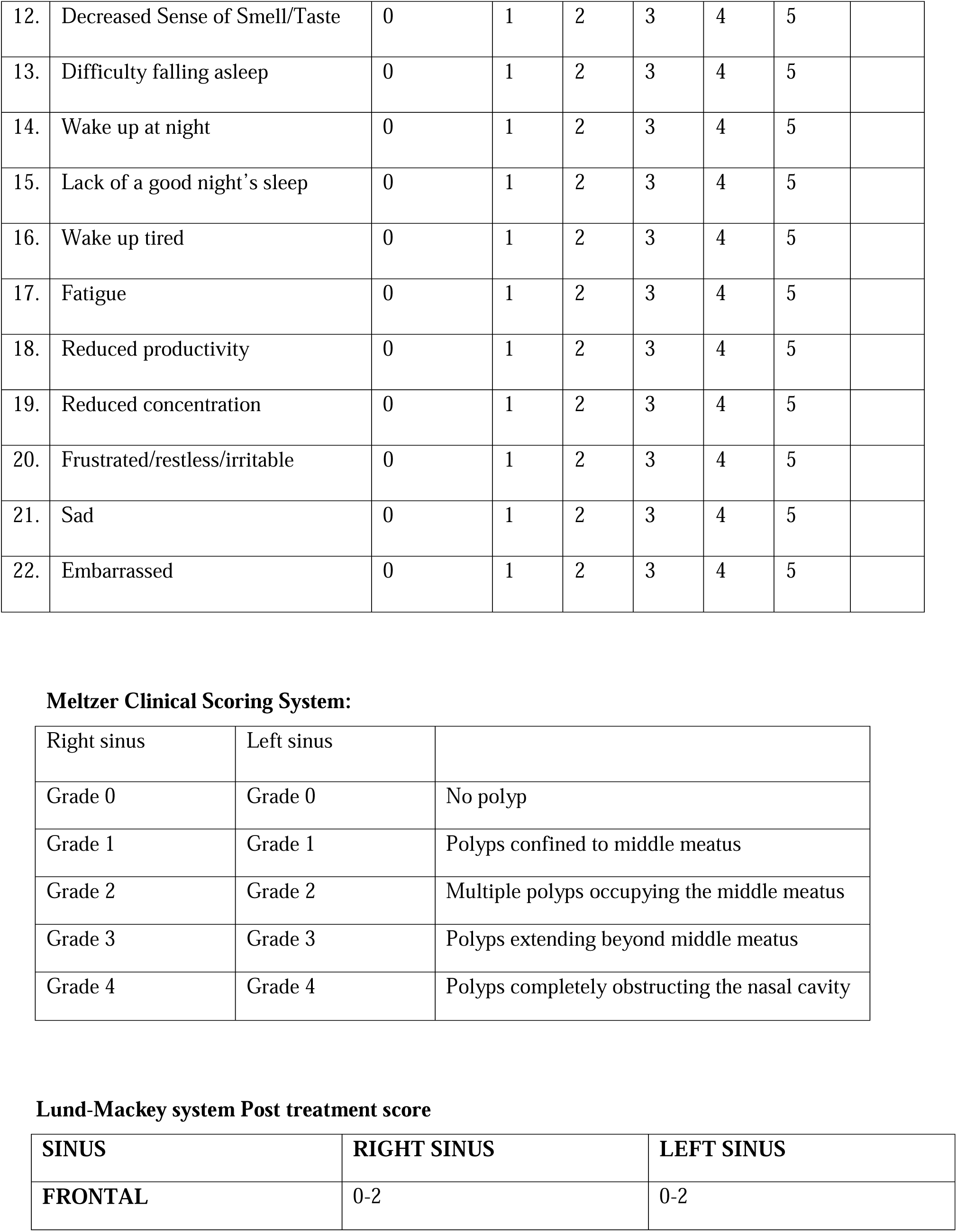
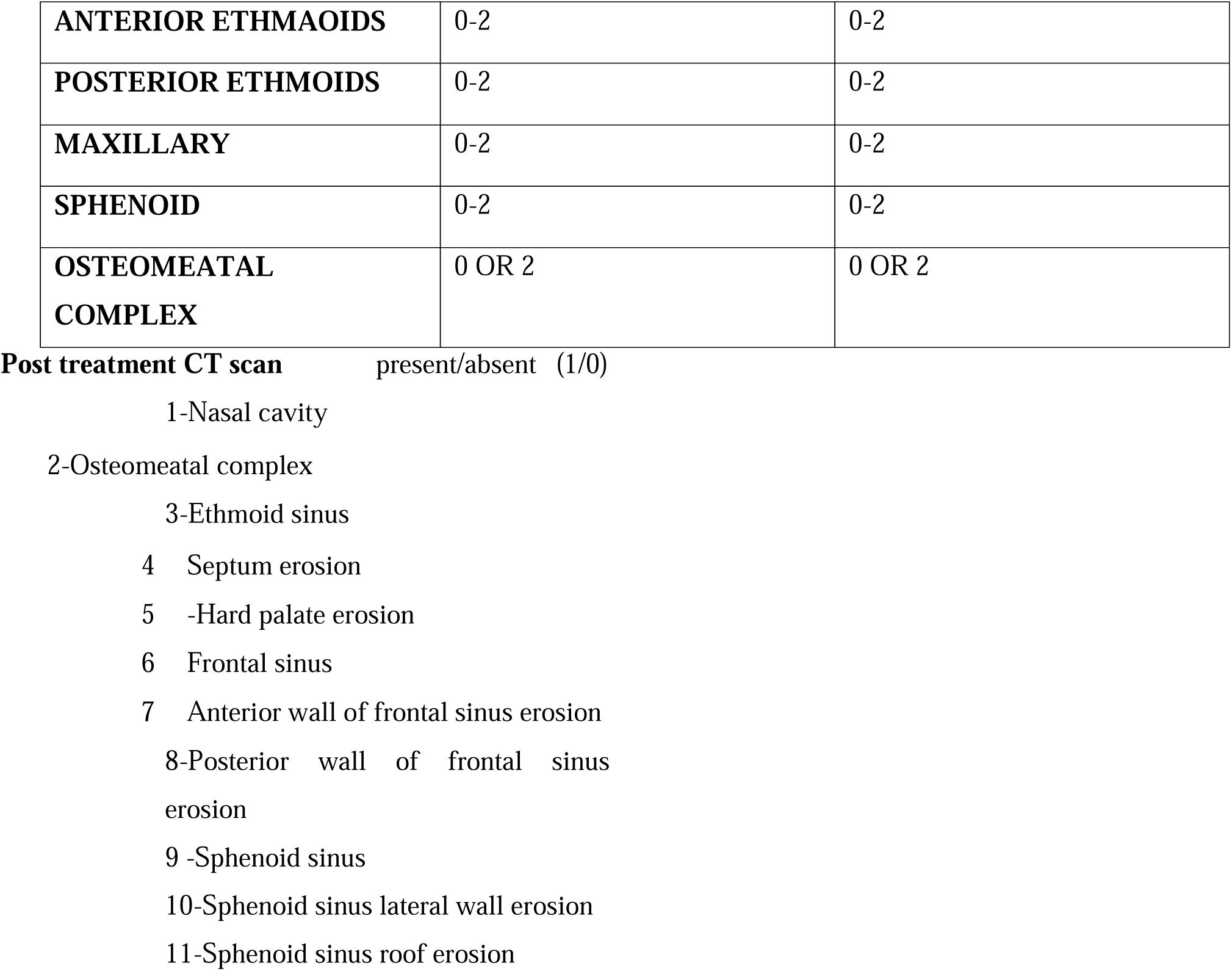
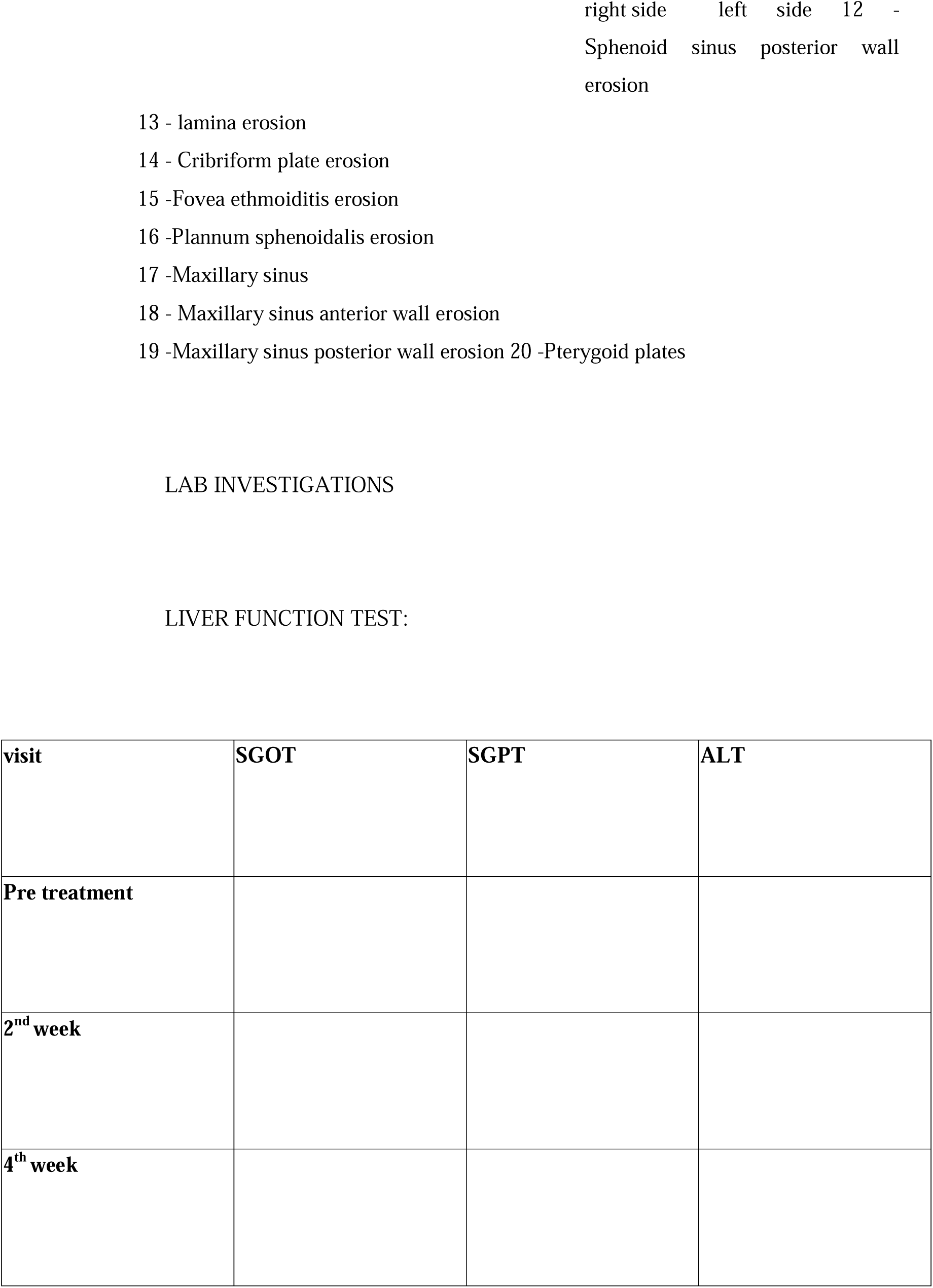
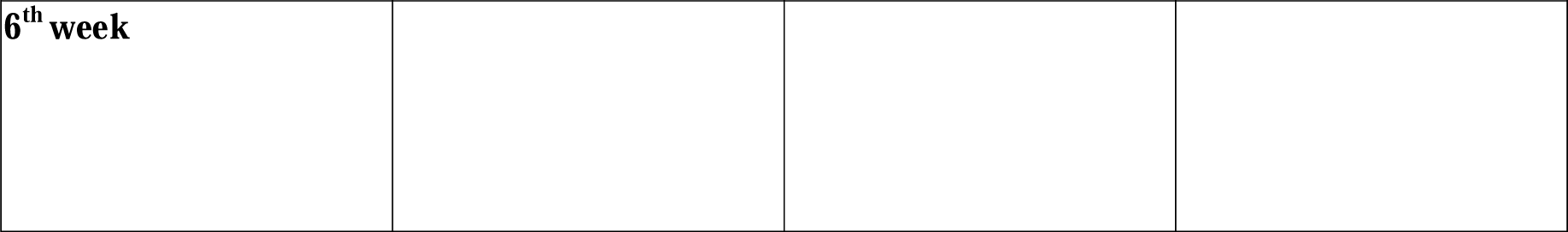

## ANNEXURE 3: CONSENT FORM IN ENGLISH

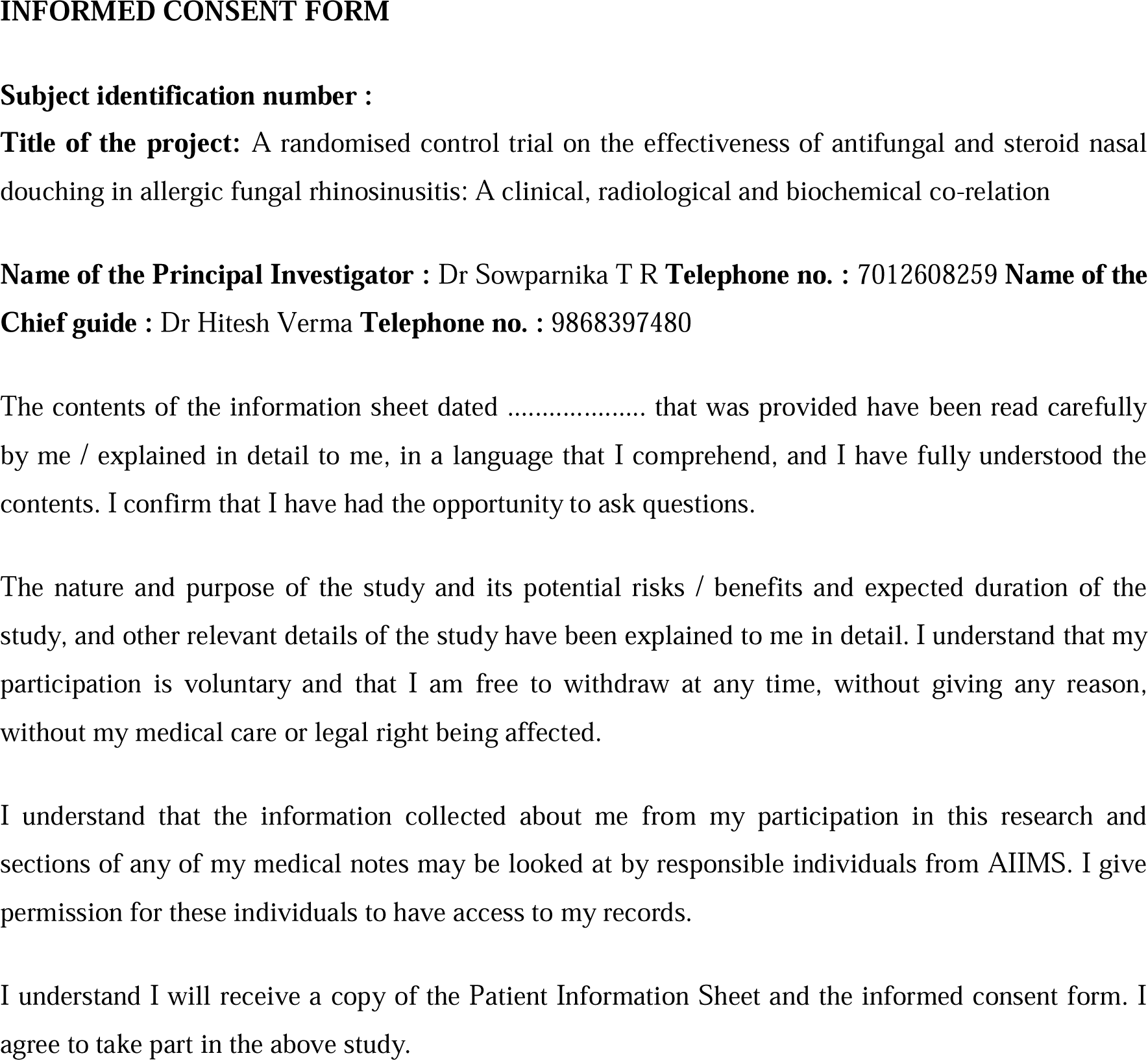
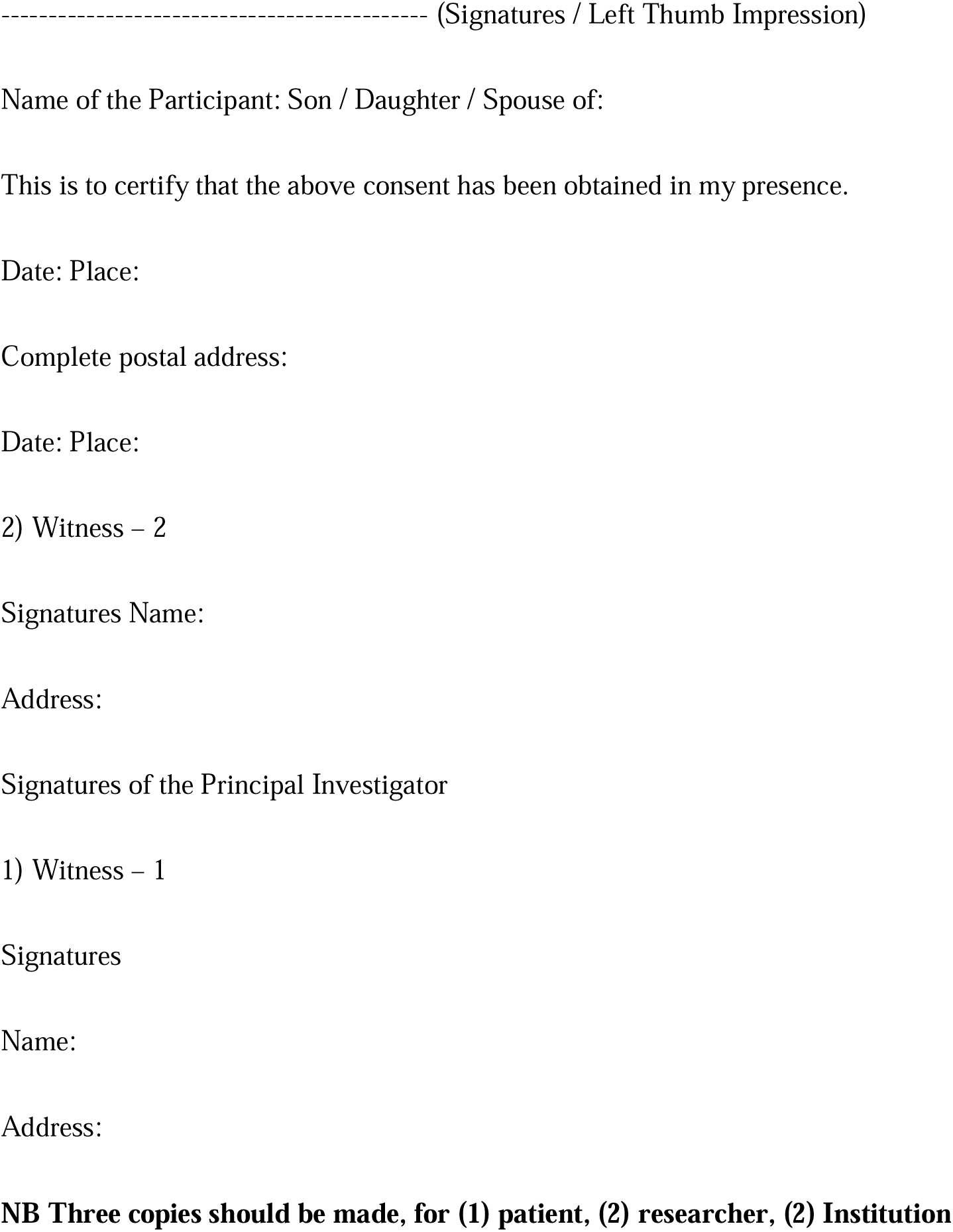

## ANNEXURE 4: CONSENT FORM IN ENGLISH

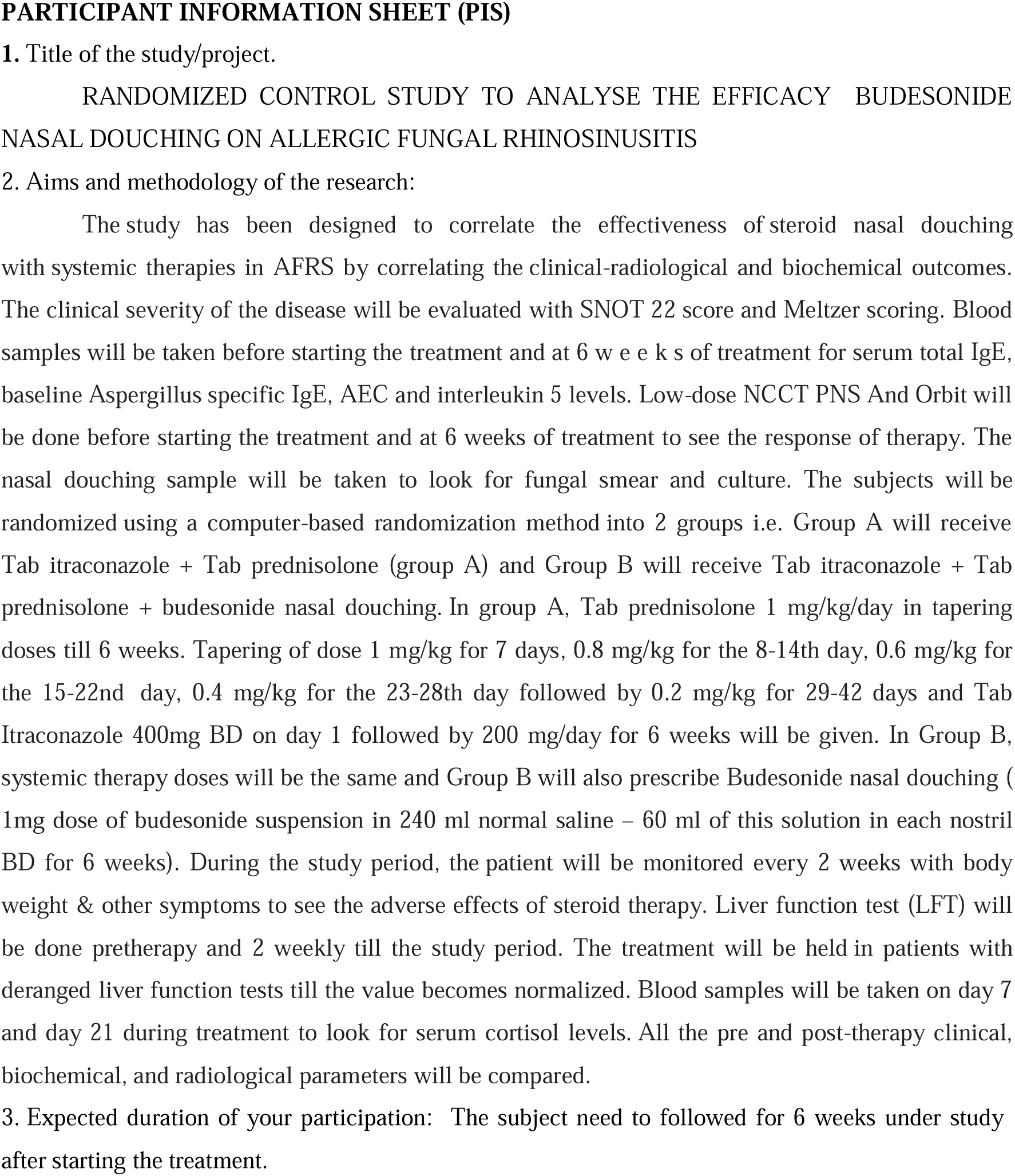
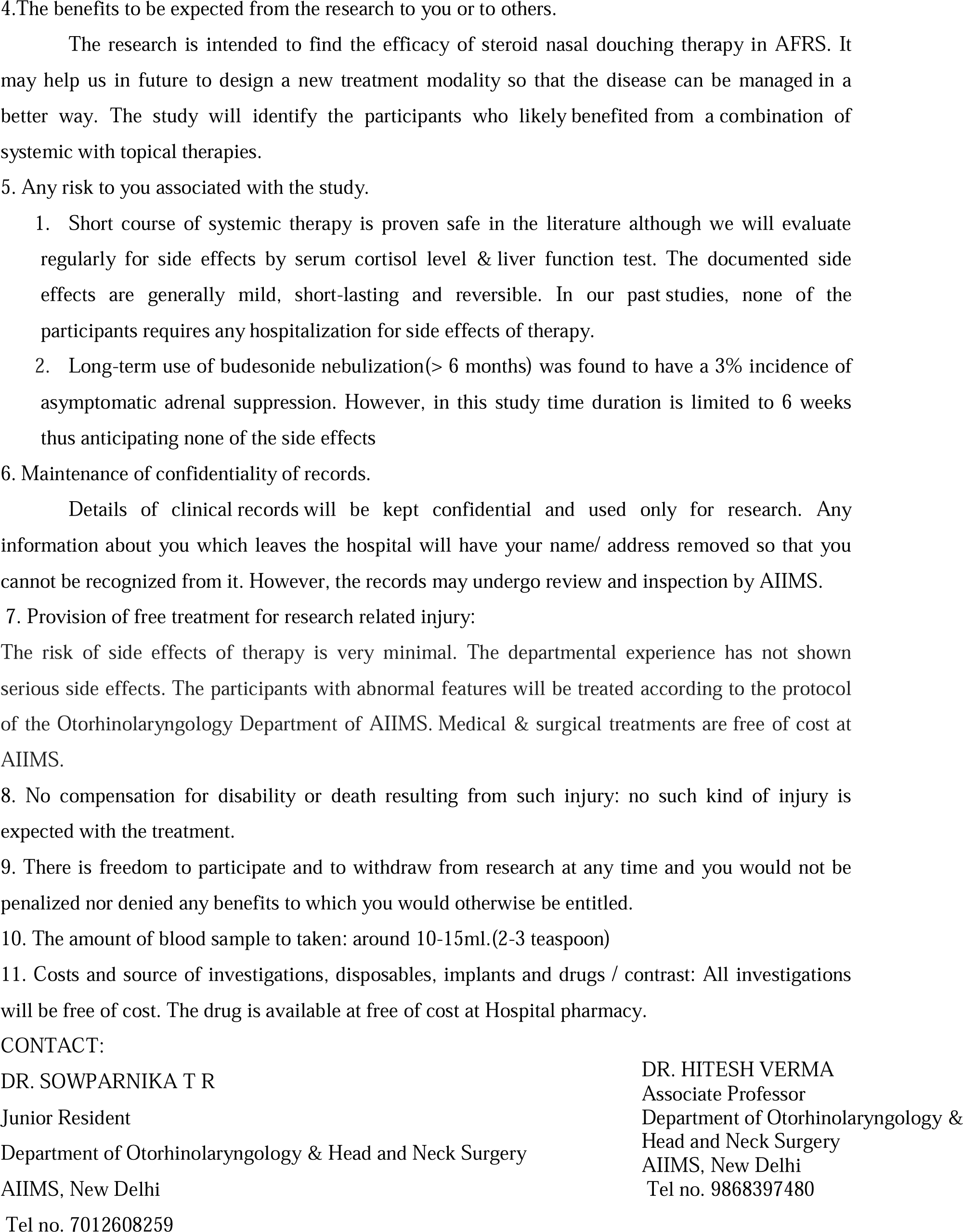

